# Development and evaluation of automated organ segmentation models for abdominal ultrasound in injured children

**DOI:** 10.64898/2026.10.27.26366648

**Authors:** Newton Addo, Kush Narang, Ritik Sinha, Naina Sabbineni, Shabir Din, Wilson Huang, Maytal Firnberg, Margaret Lin-Martore, John Mongan, Aaron Kornblith

**Affiliations:** Department of Emergency Medicine, University of California, San Francisco, CA; School of Medicine, Stanford University, Stanford, CA; University of California, Berkeley, CA; Union College, Albany Medical College, Schenectady, Albany, NY; School of Medicine, University of California, San Diego, CA; Kaiser Permanente, San Francisco, CA; Department of Pediatrics, University of California, San Francisco, CA; Department of Radiology and Biomedical Imaging, University of California, San Francisco, CA

**Keywords:** ultrasound, pediatric trauma, deep learning, image processing, multi-organ segmentation

## Abstract

**Background:** Accurate organ segmentation in pediatric focused assessment with sonography in trauma (FAST) is an important prerequisite for automated image analysis and image quality assessment, but manual segmentation is time-intensive, subject to inter- and intra-observer variability, and limits the scalability of deep learning dataset development. Although deep learning has shown promise for ultrasound organ segmentation, its application to pediatric FAST has not been evaluated.

**Objective:** To describe the development and evaluation of deep learning models to automatically localize and segment six abdominal organs in pediatric FAST.

**Methods:** We conducted a cross-sectional study using a convenience sample of archived FAST examinations from 200 injured children evaluated at a Level I pediatric trauma center. Emergency ultrasound fellowship-trained physicians manually segmented six key abdominal organs. Annotated image-mask pairs were split into training and test sets in an 80:20 ratio. We developed or fine-tuned U-Net, EVIT U-Net, UltraSAM, and nnU-Net models to automatically segment the bladder, kidney, liver, spleen, spine, and diaphragm. Segmentation accuracy was assessed using the Dice-Sørensen Coefficient (DSC) compared with manual annotations. Pediatric emergency ultrasound experts reviewed previously unlabeled FAST clips and rated the acceptability of model-generated organ masks.

**Results:** The dataset included 1,807 manually annotated image-mask pairs from 200 pediatric FAST clips. Using an 80:20 split with no overlap between video clips, 1,446 image-mask pairs were used for training and 361 for testing. Across all models and organs, mean DSC was 0.79 (95% CI: 0.75-0.84). Mean DSC was highest for the kidney and bladder, each 0.91 (95% CI: 0.87-0.95), and lowest for the diaphragm at 0.63 (95% CI: 0.60-0.67). EVIT U-Net achieved the highest DSC for kidney, liver, and spleen, performed comparably to the highest-performing models for bladder, and had a mean inference time of 54 milliseconds per image. In previously unlabeled pediatric FAST clips, experts rated model-generated organ masks as acceptable for localization in 73%-95% of clips across organs.

**Conclusion:** We describe the automatic segmentation of key abdominal organs in pediatric FAST with limited annotated data. This represents an important advancement in our image processing pipeline and can accelerate the development of large, robust datasets with segmentation for anatomy and pathology. Experts found that model-generated masks provided acceptable localization in most previously unlabeled pediatric clips, supporting the potential use of automated segmentation in pediatric FAST image analysis.

## Introduction

Pediatric trauma is a leading cause of morbidity and mortality in children worldwide,^1–3^ and the focused assessment with sonography for trauma (FAST), a point-of-care ultrasound examination in which a sequence of bedside views rapidly identifies free fluid interpreted as hemorrhage,^4^ is a key diagnostic tool in trauma care. Although FAST has been shown to decrease computed tomography usage and improve clinical outcomes in adults with blunt trauma,^3^ its utility in children is limited by variability in image acquisition and interpretation.^2,4–10^ This is especially true in technically limited studies with incomplete visualization of the hepatorenal space, splenorenal space, or pelvis, which prevents reliable exclusion of hemorrhage.^5,11,12^

Automated localization and segmentation of the abdominal organs that define these zones is essential for assessing examination quality, supporting education, and enabling automated image analysis.^4,13,14^ Currently, this process depends on manual annotation by trained experts, which is time-intensive, subject to inter- and intra-observer variability, and represents a rate-limiting step in the development of large, robust datasets for downstream deep learning applications.^15,16^

Deep learning has shown promise for automating image analysis tasks in diagnostic ultrasound, including organ segmentation, lesion detection, and view classification.^14,17,18^ Convolutional neural networks (CNNs), and the U-Net architecture in particular,^19,20^ have demonstrated strong performance across a range of medical image segmentation tasks^21^ and have been applied to abdominal ultrasound in adult populations.^22–24^ More recently, Vision Transformer (ViT)-based architectures^21,25^ and large foundation models such as Meta’s Segment Anything Model (SAM), its medical imaging adaptation (MedSAM)^26^ and its ultrasound-specific adaptation (UltraSAM),^27^ have expanded segmentation capabilities through self-attention mechanisms and large-scale pretraining on medical imaging datasets. Self-configuring frameworks such as nnU-Net have further simplified model development by automating architecture selection and hyperparameter tuning for new datasets.^28–30^ However, these architectural advances come with tradeoffs in computational resource requirements and susceptibility to overfitting on limited training data, considerations that should be relevant to clinical deployment contexts.^25^

Despite this progress, automated segmentation has not been systematically applied to pediatric FAST. Pediatric ultrasound presents unique challenges relative to adult imaging: organ size varies across age groups, anatomic planes differ, and image quality is highly heterogeneous when bedside clinicians perform studies in a fast-paced emergency environment rather than when accredited sonographers perform them under controlled conditions.^4,8,16^ Models developed on adult-only or highly curated but non-representative datasets may not generalize to this real-world variability without direct evaluation on pediatric FAST data.^17,18^

We sought to develop and evaluate automated deep learning models for organ localization and segmentation in pediatric FAST using a heterogeneous dataset of real-world ultrasound studies.

We developed and tested four architectures: U-Net, EVIT U-Net, UltraSAM, and nnU-Net, representing a range of convolutional and transformer-based approaches. We assessed both quantitative segmentation accuracy and qualitative clinical acceptability.

## Materials & Methods

### Data Sources

Three datasets were used in this study: (1) a labeled dataset of 200 FAST video clips (“FAST 200”) containing manually annotated frames for six abdominal organs, (2) an unlabeled pediatric dataset (n = 120 clips; 40 right upper quadrant [RUQ], 40 left upper quadrant [LUQ], and 40 suprapubic), and (3) an unlabeled adult dataset (n = 120 clips; same distribution by view as the unlabeled pediatric dataset). The labeled dataset was sourced from archived pediatric FAST examinations performed at UCSF Benioff Children’s Hospitals (includes one level 1 Trauma Center and one pediatric emergency department). Both pediatric labeled and unlabeled data were obtained from UCSF Benioff Children’s Hospitals. The adult dataset was obtained from an additional institution, Zuckerberg San Francisco General Hospital, a public safety-net hospital and level 1 trauma center.

FAST examinations were obtained as part of routine clinical care at the three institutions from March 21, 2018, through December 31, 2022. Each examination was performed at the point of care by treating clinicians and subsequently archived through institutional ultrasound quality assurance programs. Eligible studies included FAST examinations that were both interpreted by the performing clinician and successfully archived. All data were collected retrospectively, retrieved in raw Digital Imaging and Communications in Medicine (DICOM) format, and de-identified before analysis. The datasets were secured in a high-performance computing environment in compliance with institutional data governance standards. The University of California, San Francisco’s Institutional Review Board approved this study and waived consent for all datasets.

### Ground Truth Annotation

Within the FAST 200 dataset, pediatric emergency medicine (PEM) physicians who had completed US fellowships and trained clinical research staff manually labeled organ boundaries for six structures: bladder, kidney, liver, spleen, spine, and diaphragm. These structures were selected because they were the largest anatomic structures present in the abdominal and pelvic views of the exam. Retrospective localization of these organs can indicate transducer placement and orientation and provide information about the completeness and quality of the captured views. Annotations were performed using MD.ai (MD.ai, Inc, New York, NY)^31^ and reviewed for quality by a senior clinician.

### Data Partitioning

Annotated image-organ mask pairs were divided into training and testing sets using an 80:20 split, ensuring no video clips overlapped between datasets. Organ-specific models were trained using anatomically relevant views: the bladder model on suprapubic (sagittal or transverse) views; liver and diaphragm models on RUQ views; spleen models on LUQ views; and kidney and spine models on both RUQ and LUQ views.

### Preprocessing

All images and corresponding segmentation masks were resized to model-specific input dimensions and saved as PNG files. Pixel intensities were normalized prior to training the models. Data augmentation techniques were applied variably across all models to improve generalizability and reduce overfitting, including random horizontal and vertical flips, rotations, and cropping, with specific parameters varying nominally across model implementations.

### Model Development

We developed and evaluated four deep learning segmentation architectures that represent a range of convolutional and transformer-based approaches **(Table 1)**. All models were trained using organ-specific image-mask pairs from the FAST 200 training set and evaluated on the pediatric test data set. Model inference was performed on an AWS g5.2xlarge instance with a single NVIDIA A10G GPU. Additional details on checkpoint selection are included in the accompanying repository.

**Table 1.** Comparison of Model Implementation/features.

| **Model** | **Source** | **Implementation** | **Input image Size (px)** | **Approximate Model Size on Disk** | **Mean Inference Time\*** |
| --- | --- | --- | --- | --- | --- |
| U-Net | https://github.com/tensorflow/tensorflow | Tensorflow<br>(Trained up to 40 Epochs) | 256 x 256 | 25 MB | 35 ms |
| EVIT-UNet | https://github.com/Retinal-Research/EVIT-UNET | Pytorch<br>(Trained up to 300 Epochs) | 224 x 224 | 250 MB | 54 ms |
| UltraSam | https://github.com/CAMMA-public/UltraSam | Pytorch<br>(Trained up to 2000 Epochs) | 1024 x 768<br>(padded) | 360 MB | 124 ms |
| nnU-Net | https://github.com/MIC-DKFZ/nnUNet | Pytorch<br>(Trained up to 250 Epochs) | 960 x 720 | 350 MB | 172 ms |

#### U-Net

U-Net, a fully convolutional network with a symmetric encoder-decoder architecture, served as a baseline model given its established performance in medical image segmentation and localization tasks.^19^ This architecture is well-suited to datasets of limited size when sufficient data augmentation is applied. The U-Net model was implemented in TensorFlow and trained for up to 40 epochs with an input image size of 256×256 pixels.

#### EVIT U-Net

EVIT U-Net extends the U-Net encoder-decoder architecture by incorporating an efficient Vision Transformer (ViT)-based self-attention mechanism,^20^ preserving the spatial symmetry of U-Net while reducing the computational overhead typically associated with full-scale transformer models. This architecture was implemented in PyTorch and trained for up to 300 epochs with an input image size of 224×224 pixels.

#### UltraSAM

UltraSAM is based on Meta’s Segment Anything Model (SAM)^32^ and incorporates a ViT encoder fine-tuned on a large ultrasound-specific dataset of 280,000 image-mask pairs (US-43),^27^ making it an ultrasound-adapted foundation model for downstream segmentation tasks. This model was implemented in PyTorch and trained for up to 2,000 epochs with padded input images of 1,024×768 pixels.

#### nnU-Net

NnU-Net is a self-configuring segmentation framework that automatically configures the network and training to the characteristics of the input dataset.^29,30^ This model was implemented in PyTorch and trained for up to 250 epochs with an input image size of 960×720 pixels.

### Evaluation

#### Segmentation Accuracy

Model performance was assessed on the test set using the Dice-Sørensen Coefficient (DSC), calculated for each organ independently at the best model checkpoint. DSC ranges from 0 to 1, with higher values indicating greater overlap between predicted and ground truth masks.

#### Clinical Acceptability

Following the selection of a single model for implementation, we assessed the clinical acceptability of model-generated organ masks on the two previously unlabeled datasets. **(Figure 1)** EVIT U-Net model-generated organ masks were created and applied to all frames from 120 pediatric and 120 adult clips. Predicted masks were then reviewed by four PEM ultrasound experts for the middle 60 frames of each clip using the MD.ai image review platform **(Figure 1)**. For each clip, one reviewer assessed (1) whether the model achieved acceptable organ localization, and (2) whether the image quality for the included clip was acceptable for clinical interpretation. Results are reported as the proportion of clips rated acceptable out of 40 reviewed per view per cohort (**Table 3**).

**Figure 1.**
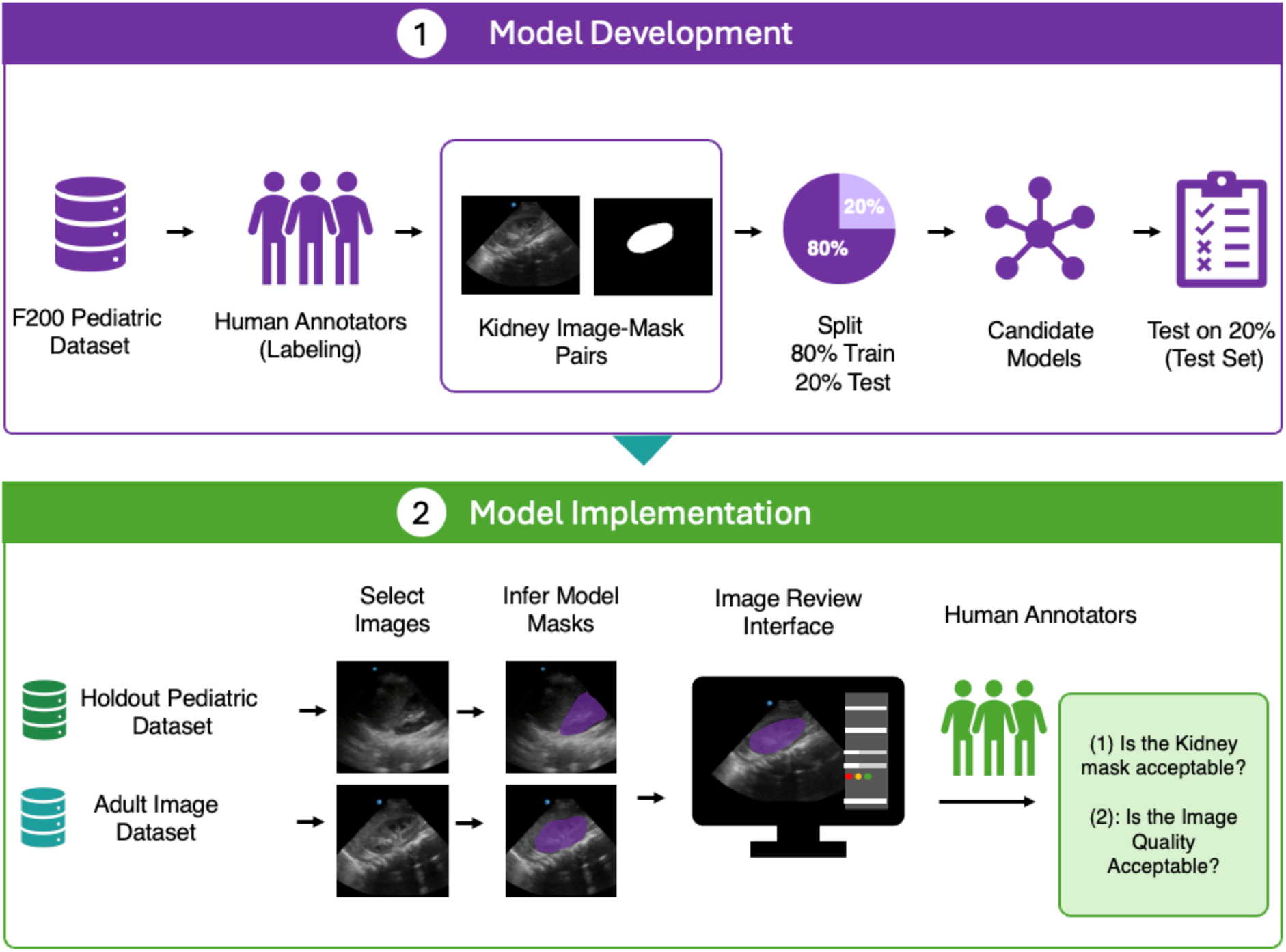
Study Flow. Overview of the study workflow for model development and clinical evaluation. Manually annotated FAST images were divided into training (80%) and test sets (20%) with no overlap between video clips. Candidate segmentation models were trained using organ-specific image-mask pairs and evaluated on the held-out pediatric test set. Following model selection, EVIT U-net masks were applied to previously unlabeled pediatric and adult FAST clips. Pediatric emergency ultrasound experts reviewed the resulting model-generated masks for acceptable organ localization and assessed whether clip image quality was acceptable for clinical interpretation.

## Results

The labeled dataset comprised 1,817 manually annotated image-mask pairs across six organ targets: bladder (n=344), kidney (n=306), liver (n=445), spleen (n=172), spine (n=273), and diaphragm (n=267). The median age of the sample in the derivation and testing sets was 10 (IQR: 5, 14), and 56% were male. Full characteristics of the sample can be found in the **online supplemental Table 1.** Following an 80:20 split, approximately 1,446 image-mask pairs were used for training and 361 for testing, with no overlap of video clips across datasets.

Representative machine-generated organ segments are shown in **Figure 2**.

**Figure 2.**
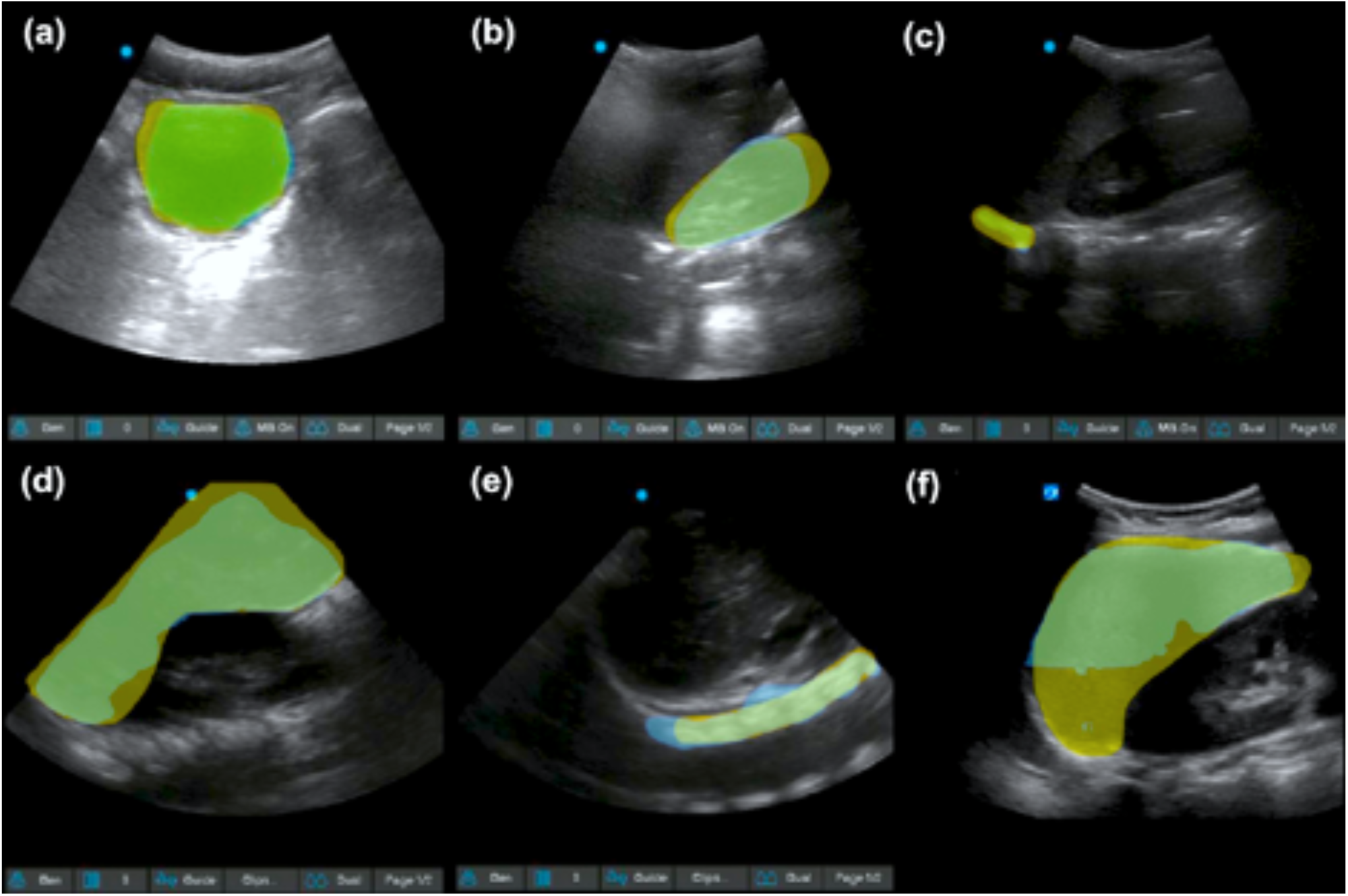
Machine-generated organ segmentation of FAST. Representative machine-generated and manually annotated segmentation for six organs visualized during pediatric FAST: (a) bladder, (b) kidney, (c) diaphragm, (d) spleen, (e) spine, (f) liver. Machine-generated segmentation is shown in blue, manual segmentation in yellow, and regions of overlap between the machine-generated and manual segmentation in green.

### Segmentation Accuracy

All four models achieved moderately high segmentation accuracy across the six organ targets **(Table 2**, **Figure 3)**. Performance was highest for the kidney and bladder, and lowest for the diaphragm and spine across all models. EVIT U-Net achieved the highest DSC for kidney (0.93), liver (0.90), and spleen (0.82), and was comparable to UltraSAM for bladder (0.92 vs. 0.91) and liver (0.90 vs. 0.90). U-Net performed similarly to EVIT U-Net for kidney (0.92 vs. 0.93) and bladder (0.90 vs. 0.92), but showed lower performance for spleen (0.70 vs. 0.82). nnU-Net demonstrated the lowest DSC for the spleen (0.60), despite achieving competitive scores for the bladder (0.92) and liver (0.89). The diaphragm was the lowest-performing target across all models (DSC range: 0.61-0.67), followed by the spine (DSC range: 0.66-0.70). EVIT U-Net was selected for implementation in the clinical review interface based on its overall segmentation performance across organ targets.

**Figure 3.**
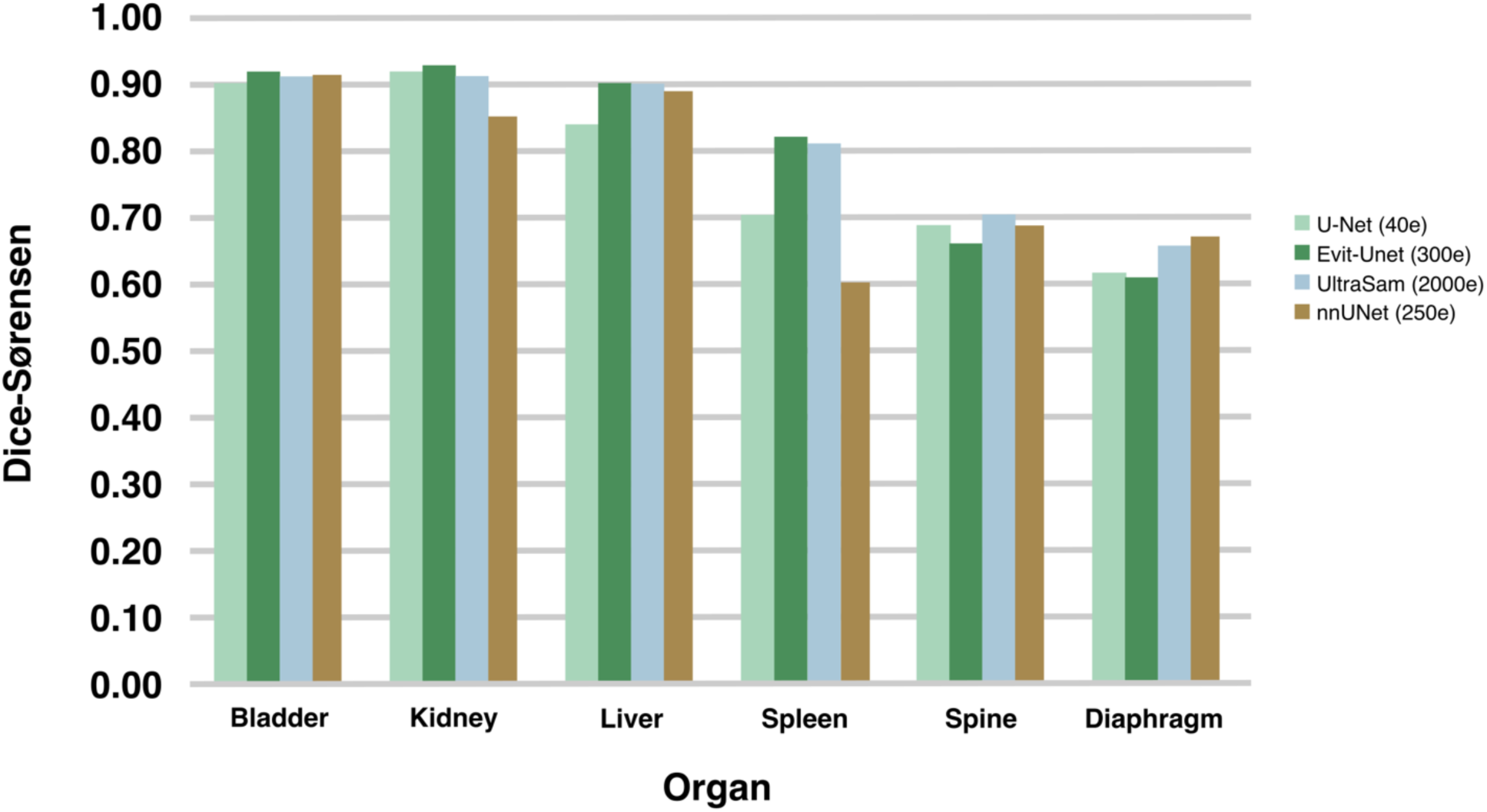
**Dice-Sørensen similarity by model and organ**. Segmentation performance of U-Net, Evit-Unet, UltraSAM, and nnU-Net across six organ targets in the pediatric FAST. The Dice-Sørensen Coefficient (DSC) was used to assess performance, with higher values indicating greater overlap between model-generated and manually annotated organ segmentation masks.

**Table 2.** Model segment Dice-Sørensen (DSC) similarity on pediatric Focused Assessment with Sonography for Trauma (FAST) test set.

| **FAST Organ** | **Bladder** | **Kidney** | **Liver** | **Spleen** | **Spine** | **Diaphragm** |
| --- | --- | --- | --- | --- | --- | --- |
| Manually Labeled Images | n=344 | n=306 | n=445 | n=172 | n=273 | n=267 |
| **Model** |  |  |  |  |  |  |
| U-Net | 0.90 | 0.92 | 0.84 | 0.70 | 0.69 | 0.62 |
| EVIT-UNet | 0.92 | 0.93 | 0.90 | 0.82 | 0.66 | 0.61 |
| UltraSAM | 0.91 | 0.91 | 0.90 | 0.81 | 0.70 | 0.66 |
| nnU-Net | 0.92 | 0.85 | 0.89 | 0.60 | 0.69 | 0.67 |

Mean inference time per image increased substantially with model complexity, ranging from 35 milliseconds (ms) for U-Net to 172 ms for nnU-Net. EVIT U-Net and UltraSAM required 54 ms and 124 ms per image, respectively. Average model size on disk showed a similar pattern, from 25 megabytes (MB) to 360 MB (UltraSAM), with EVIT U-Net at 250 MB **(Table 1)**.

### Clinical Acceptability

EVIT U-Net was applied to all 120 previously unlabeled pediatric clips and 120 adult clips. Expert reviewers assessed the model-generated organ masks for acceptable localization and acceptable image quality across RUQ, LUQ, and pelvic views in both cohorts **(Table 3)**.

**Table 3.**
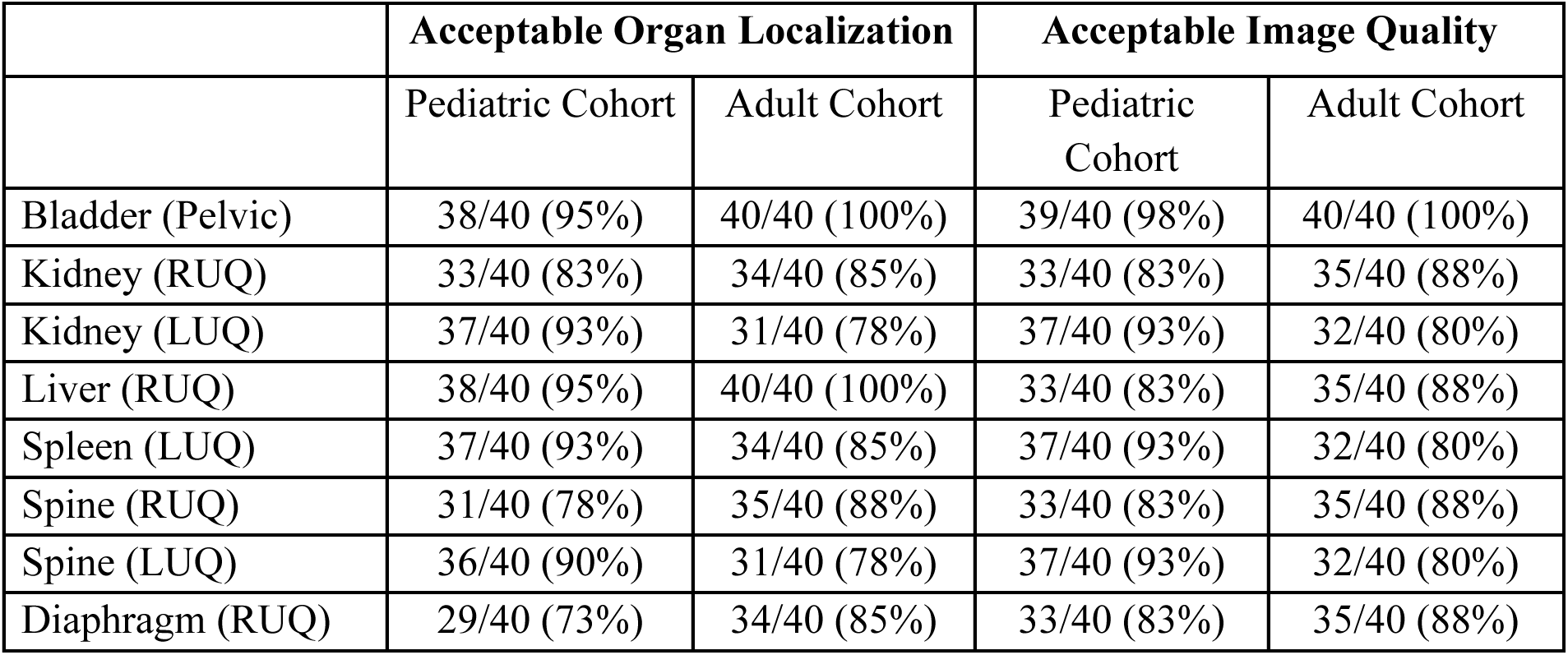
Expert assessment of unlabeled images and predicted masks.

|  | Acceptable Organ Localization |  | Acceptable Image Quality |  |
| --- | --- | --- | --- | --- |
|  | Pediatric Cohort | Adult Cohort | Pediatric Cohort | Adult Cohort |
| Bladder (Pelvic) | 38/40 (95%) | 40/40 (100%) | 39/40 (98%) | 40/40 (100%) |
| Kidney (RUQ) | 33/40 (83%) | 34/40 (85%) | 33/40 (83%) | 35/40 (88%) |
| Kidney (LUQ) | 37/40 (93%) | 31/40 (78%) | 37/40 (93%) | 32/40 (80%) |
| Liver (RUQ) | 38/40 (95%) | 40/40 (100%) | 33/40 (83%) | 35/40 (88%) |
| Spleen (LUQ) | 37/40 (93%) | 34/40 (85%) | 37/40 (93%) | 32/40 (80%) |
| Spine (RUQ) | 31/40 (78%) | 35/40 (88%) | 33/40 (83%) | 35/40 (88%) |
| Spine (LUQ) | 36/40 (90%) | 31/40 (78%) | 37/40 (93%) | 32/40 (80%) |
| Diaphragm (RUQ) | 29/40 (73%) | 34/40 (85%) | 33/40 (83%) | 35/40 (88%) |

Acceptable localization rates ranged from 73% to 95% in the pediatric cohort and from 78% to 100% in the adult cohort. Performance was highest for the bladder (pediatric: 95%, adult: 100%) and liver (pediatric: 95%, adult: 100%), and lowest for the diaphragm (pediatric: 73%, adult: 85%) and kidney on RUQ views (pediatric: 83%, adult: 85%). Acceptable image quality ratings ranged from 83% to 98% in the pediatric cohort and from 80% to 100% in the adult cohort.

Bladder segments again achieved the highest image quality ratings in both cohorts (pediatric: 98%, adult: 100%), while spleen, kidney on LUQ views, and spine on LUQ views showed lower image quality ratings in the adult cohort (80% each).

## Discussion

We trained and evaluated four deep learning models for automated organ localization and segmentation in pediatric FAST, demonstrating moderately high segmentation accuracy across six abdominal organ targets and clinically acceptable performance on previously unlabeled pediatric and adult datasets. To our knowledge, this is the first systematic comparison of convolutional and transformer-based segmentation architectures applied to real-world pediatric FAST, and the first to evaluate model-generated organ masks for clinical acceptability in this population.

Performance was most consistent for the kidney and bladder (DSC: 0.91–0.93 and 0.90–0.92, respectively), likely reflecting the relatively distinct echogenic boundaries of these organs. The diaphragm and spine were the most challenging targets, consistent with prior reports of difficulty segmenting thin, curvilinear, or acoustically complex structures in ultrasound.^13,33,34^ Segmentation failures were likely to occur when the original image was identified as being of poor image quality rather than due to systematic model error, suggesting that upstream image-quality filtering may substantially improve deployed performance.

EVIT U-Net achieved a high overall segmentation performance and was selected for implementation. We found that its demonstrated performance was consistent with the hypothesis that incorporating self-attention mechanisms into a U-Net backbone improves the model’s ability to capture long-range spatial dependencies^35^ without the computational overhead and risk of overfitting associated with a full-scale transformer.^20,25^

A central finding of this study is the relationship between model complexity, computational resource requirements, and segmentation performance. The four models span a clear spectrum: U-Net is the simplest and fastest (35 ms mean inference time, 25 MB on disk), while UltraSAM is the largest and slowest (124 ms, 360 MB), with EVIT U-Net and nnU-Net at intermediate positions. Notably, segmentation performance did not scale linearly with model complexity.

EVIT U-Net, the second-smallest model, achieved comparable or superior accuracy to both UltraSAM and nnU-Net on most organ targets while requiring substantially fewer computational resources and shorter inference times. The diminishing returns observed in larger models could be attributable, in part, to overfitting on a training dataset of limited size, in which the additional parameters of UltraSAM and nnU-Net may not be fully leveraged. Not overlooked are differences in the model training pipeline, such as native input resolution and the number of training epochs, which could also be sources of confounding in segmentation performance.

We also found limited performance benefit from ultrasound-specific pretraining. UltraSAM, despite being fine-tuned on 280,000 ultrasound-specific image-mask pairs, did not outperform EVIT U-Net on most organ targets. This is consistent with emerging evidence that domain-specific pretraining does not always confer downstream advantages when the target dataset is small and heterogeneous, and additionally suggests that architecture and training strategy may matter more than the pretraining domain for such tasks.^37^ A direct comparison of baseline SAM, MedSAM, and UltraSAM in this setting would be a valuable future extension.

Expert reviewers found model-generated organ masks to be acceptable in most reviewed clips, with localization rates ranging from 73-95% in the pediatric cohort and 78-100% in the adult cohort. Using the number of clips with only acceptable image quality as the denominator further increases the proportion of model-generated masks rated acceptable over 90% in all organs. In this way, these findings, along with their agreement with the DSC values, provide complementary quantitative and qualitative evidence. The finding that acceptability rates were comparable between pediatric and adult cohorts, despite the adult data being entirely out-of-distribution relative to the training set, suggests some amount of generalizability and supports the potential for broader application of this approach across trauma settings.

This study has several limitations. First, the labeled training dataset was limited to 200 FAST video clips from only two centers, constraining the diversity of imaging conditions, patient demographics, and operator techniques in model training. Second, acceptability was evaluated on unlabeled data from three centers, but this does not constitute comprehensive external validation against ground truth annotations. The acceptability analysis was performed only on EVIT U-Net; a parallel review across four architectures would enable a more complete comparison. However, in this context, we consider all four architectures suitable choices for similar segmentation tasks. Third, results should not be generalized beyond the included datasets and imaging contexts without further validation.

## Conclusion

We developed and evaluated an automated segmentation pipeline for key abdominal organs in pediatric FAST using a limited annotated dataset. This represents an important advancement in our image processing pipeline and will accelerate the development of large, robust datasets with segmentation for anatomy and pathology. Experts found that model-generated masks provided acceptable localization in most previously unlabeled pediatric clips, supporting the potential use of automated segmentation in pediatric FAST image analysis and annotation workflow.

Automated organ segmentation could substantially reduce the time and variability associated with manual annotation, enabling the construction of larger, more heterogeneous training datasets for future multistage models targeting free fluid detection, view quality assessment, and technically limited study identification. By evaluating and sharing this approach, we aim to provide a tool that investigators can use to generate model-assisted segmentation masks on their own ultrasound datasets. We hope this framework will enable scalable, standardized data annotation and accelerate collaborative development and validation of AI tools for ultrasound.

## Acknowledgments

The authors acknowledge the UCSF Bakar Computational Health Sciences Institute and the UCSF Center for Intelligent Imaging for their support and for providing an environment that promotes collaboration at the intersection of medical imaging, artificial intelligence, and clinical care.

## Conflict of Interest

Aaron Kornblith and Newton Addo are consultants and co-founders of the University of California, San Francisco spinout CaptureDx, Inc. The entity had no role in the design, planning, or execution of the study. Aaron Kornblith is supported by the Eunice Kennedy Shriver National Institute of Child Health and Human Development of the National Institutes of Health under award number K23HD110716 (AK). John Mongan receives royalties or licenses from GE Healthcare through UCSF and research funding from SYRMOUNT and the NIH. This information, content, and conclusions are those of the authors. They should not be construed as the official position or policy of, nor should any endorsements be inferred by HRSA, HHS, or the US Government. The other authors have no conflicts of interest to report.

## Data Sharing

*Model checkpoints and code for model training and evaluation will be publicly available at* https://github.com/Kornblith-Lab/US-AbdomenSeg*. Patient-level data and images will not be distributed with this repository*.

## Abbreviations

U-Net: U-Network
EVIT: Efficient Vision Transformer
FAST: Focused Assessment with Sonography in Trauma
UltraSAM: Ultra Segment Anything Model
DSC: Dice-Sørensen Coefficient
RUQ: right upper quadrant
LUQ: left upper quadrant

## Notes

### Author Declarations

The IRB of the University of California, San Francisco gave ethical approval for this work.

